# High Frequency and Prevalence of Community-Based Asymptomatic SARS-CoV-2 Infection

**DOI:** 10.1101/2020.12.09.20246249

**Authors:** Lao-Tzu Allan-Blitz, Isaac Turner, Fred Hertlein, Jeffrey D. Klausner

## Abstract

Approximately 20-40% of SARS-CoV-2 infection is asymptomatic; however, data are limited on drivers of such infection. Among over 730,000 SARS-CoV-2 test results in Los Angeles between August-October, 2020, we found heterogenous frequencies of asymptomatic infection among various sup-populations. Further research is needed to delineate drivers of asymptomatic SARS-CoV-2 infection.

## Background

The economic impact of blanket closures in the wake of the SARS-CoV-2 pandemic will be felt for years to come (1). Among other instances of infectious disease outbreaks, highly targeted interventions are feasible in part because we identify specific exposures that place individuals at increased risk for infection. If we are able to understand the exposures that drive the SARS-CoV-2 pandemic, we may be able to mitigate the economic impact by facilitating narrower public health interventions.

Of particular interest are the exposures that result in asymptomatic infection, which account for approximately 20-40% of infections (2) and contribute significantly to the continued transmission of SARS-CoV-2 (3). Some studies suggest that younger age and lack of other comorbidities may be particularly associated with asymptomatic SARS-CoV-2 infection, however more detailed data are lacking (4). Given that the spread of the SARS-CoV-2 pandemic appears to be heterogeneous (5), concentrating within specific hotspots of localized spread, developing a thorough understanding of the drivers of asymptomatic infection may be instrumental in the fight against the pandemic. We thus aimed to describe the frequency and prevalence of asymptomatic infection among a community-based sample in Los Angeles.

## Methods

We evaluated SARS-CoV-2 RNA test results from a large testing program in Los Angeles between August-October 2020. Individuals presented to testing and were asked via a confidential online survey if in the last 14 days they had been contacted by local public health authorities about a known SARS-CoV-2 exposure, they visited any of a list of public places, or they spent time with five or more strangers. We collected those data, as well as demographic data (age, gender, race, ethnicity, place of employment) and report of any symptoms from a pre-specified list. We then conducted a cross-sectional analysis to determine the frequency of infection among asymptomatic and symptomatic individuals.

The Mass General Brigham institutional review board deemed the analysis of de-identified data did not constitute human subjects’ research (2020P003530).

## Findings

We analyzed more than 730,000 test results (see Table), of which 54.4% were among women and 41.6% reported Hispanic or Spanish ethnicity. The mean age was 34 years. The prevalence of SARS-CoV-2 infection among asymptomatic individuals was 4.2%. Among all who tested positive, 42.3% were asymptomatic. We found a higher prevalence of asymptomatic infection among individuals who reported work in construction and among racial and ethnic minorities. We also identified a higher prevalence of asymptomatic infection among individuals who had been contacted by a representative from a local health department regarding a known SARS-CoV-2 exposure.

**Table :** Prevalence of SARS-CoV-2 Positivity Among Symptomatic and Asymptomatic Individuals Presenting for Testing in Los Angeles, August - October 2020

|  |  | Individuals Tested for SARS-CoV-2 |  |
| --- | --- | --- | --- |
|  |  | (n=730,801) |  |
|  |  | Asymptomatic<br>(n=402,918) | Symptomatic<br>(n=289,052) |
| **Total** |  | 4.2% | 8.0% |
| **Age** |  |  |  |
|  | <18 years (n=56,127) | 9.5% | 14.4% |
|  | 18-24 years (n=115,608) | 4.9% | 8.8% |
|  | 25-34 years (n=263,306) | 2.8% | 5.8% |
|  | 35-49 years (n=180,729) | 3.6% | 7.8% |
|  | 50-64 years (n=89,322) | 4.7% | 10.1% |
|  | ≥ 65 years (n=25,709) | 4.1% | 9.3% |
| **Gender** |  |  |  |
|  | Female (n=397,159) | 3.9% | 7.3% |
|  | Male (n=330,269) | 4.6% | 9.0% |
|  | Other (n=3,373) | 1.6% | 2.5% |
| **Race** |  |  |  |
|  | American Indian or Alaska Native (n=3,936) | 5.8% | 10.3% |
|  | Black or African American (n=34,998) | 2.2% | 5.1% |
|  | Native Hawaiian or Other Pacific Island (n=5,200) | 2.0% | 5.0% |
|  | Prefer not to share (n=69,888) | 6.1% | 11.0% |
|  | Asian (n=82,480) | 1.0% | 2.5% |
|  | Multiracial (n=37,792) | 1.8% | 4.4% |
|  | Other (n=192,255) | 9.4% | 14.9% |
|  | White (n=304,252) | 2.3% | 4.8% |
| **Ethnicity** |  |  |  |
|  | Hispanic (n=304,185) | 9.3% | 13.8% |
|  | Non-Hispanic (n=399,623) | 1.4% | 3.1% |
| **Month** |  |  |  |
|  | August (n=273,198) | 5.7% | 10.1% |
|  | September (n=235,854) | 3.5% | 6.5% |
|  | October (n=221,749) | 3.3% | 6.5% |
| **Employment\*** |  |  |  |
|  | Construction Worker (n=529) | 5.7% | 10.1% |
|  | Delivery, Transportation, Ride-Share (n=1,639) | 2.3% | 5.4% |
|  | First Responder (n=12,043) | 2.6% | 7.4% |
|  | Government Employee (n=716) | 1.7% | 7.4% |
|  | Grocery Store Worker (n=8,387) | 3.1% | 7.5% |
|  | Healthcare Personnel (n=56,174) | 3.4% | 6.4% |
|  | Media Employee (584) | 1.8% | 4.9% |
|  | Correctional Worker (n=131) | 0.0% | 12.5% |
|  | Disability Care Provider (n=5,918) | 1.6% | 3.7% |
|  | Education (n=3,771) | 1.6% | 4.8% |
|  | Elderly Care (n=1,862) | 2.9% | 5.5% |
|  | Food Services (n=53,372) | 3.8% | 7.8% |
|  | Retail or Manufacturing (n=1,809) | 2.5% | 5.7% |
|  | None of the Above (n=54,174) | 3.0% | 5.6% |
| **Notified About a Positive Contact in Last 14 days\*†** |  |  |  |
|  | No (n=25,556) | 2.1% | 4.1% |
|  | Yes (n=140,858) | 9.6% | 15.3% |
| **Visited Public Places in Last 14 Days\*** |  |  |  |
|  | Yes (n=90,087) | 0.9% | 3.4% |
| **Spent time with $\geq 5$ Strangers in Last 14 Days\*** |  |  |  |
|  | Yes (n=3,765) | 2.3% | 6.4% |

\* Data collection methods changed to include these questions as of September 15th 2020
† Contacted by local health department via contact-notification efforts

## Discussion

We identified a high proportion of asymptomatic infection among those who tested positive for SARS-CoV-2. Additionally, the prevalence of infection among the asymptomatic fraction was high, providing support for screening of asymptomatic individuals at risk for infection. The increased prevalence of infection (asymptomatic and symptomatic) among those who reported being contacted by a representative from a local health department about a known SARS-CoV-2 exposure provides strong evidence of the benefits of contact-notification in case-identification.

The different frequencies of asymptomatic infection among the different employment categories likely represent heterogeneity in risk of exposure. However, further research into particular exposures within different occupations may be of use in delineating the drivers of asymptomatic infection. Our data were limited in that we did not have more exposure data related to dining or social behaviors.

Thus, testing centers should strive to routinely collect data on potential exposures in the past 7 days. Data regarding specific venues of exposure (e.g. gyms, places of worship, parks, museums, hospitals, nursing homes, overnight camps, hotels, beaches, movie theaters, bars, restaurants, and airports) among individuals who test both positive and negative will enable more precise delineation of recent transmission events in real time. Such geographic information may function to enhance network mobility data to identify recent hotspots and venues where transmission mitigation efforts would be most helpful (6). More granular exposure data might enable public health officials to move from broad county and state lockdowns towards a more, targeted less harmful approach. The identification of particular high-risk activities or business practices might allow regulators like state occupational safety and health administrations to propose or enforce regulations to reduce the spread of infection.

Given the relatively low prevalence of infection in certain areas there is ample opportunity to enhance prevention of SARS-CoV-2 spread through the use of exposure data and strategic evidence-based interventions. In the meantime, continued screening of at-risk persons regardless of symptom status is warranted.

## Data Availability

The data is not available at this time

## Acknowledgements

The authors would like to acknowledge Curative Inc. and the City of Los Angeles.

